# Characterization of *APOE* Christchurch carriers in 455,306 UK Biobank participants

**DOI:** 10.1101/2022.12.05.22283048

**Authors:** Karen Y. He, Ekaterina A. Khramtsova, Alfredo Cabrera-Socorro, Yanfei Zhang, Shuwei Li, Brice A. J. Sarver, Bart Smets, Qingqin S. Li, Louis De Muynck, Antonio R. Parrado, Simon Lovestone, Mary Helen Black

**Author notes:** These authors contributed equally to this work (Karen Y. He, Ekaterina A. Khramtsova).

## Abstract

**Importance:** A few case studies have reported the APOE Christchurch (APOECh) variant to confer protective effects for Alzheimer’s disease (AD) and a higher risk of premature cardiovascular disease (CVD). However, these studies primarily focused on a single individual or siblings from the same family.

**Objective:** We sought to characterize the clinical characteristics of individuals with APOECh variation in 455,306 participants of the UK Biobank (UKB) to determine whether it is associated with AD protection or cardiovascular risk.

**Design, Setting, and Participants:** A total of 37 individuals were identified as heterozygous carriers of APOECh in UKB sequencing data as of March 2022, resulting in allele frequency consistent with gnomAD (4.06×10^−5^). We limited our study to 36 European carriers and generated a noncarrier cohort matched on age, sex, and ancestry. Case-control analyses were performed to evaluate the frequency of 11 binary traits and differences in distributions of 80 quantitative traits and 10 polygenic risk scores (PRS) for lipid traits, CVD, and neurodegenerative diseases.

**Main Outcomes and Measures:** We compared the frequencies of binary traits (binomial distribution probability) and distributions of quantitative traits and PRS (Kolmogorov-Smirnov test).

**Results:** All 37 carriers are free of AD and only 4 have a parental history of AD. There are 22 out of 37 carriers with ≥1 cardiovascular (CV) condition in clinical and/or self-reported data, two of whom passed away due to heart disease. However, frequency of CVD, dyslipidemia, and hypertension is not enriched in carriers compared with matched non-carriers. Additionally, apolipoprotein B (apoB) is significantly lower in APOECh carriers before (p=0.004) and after statin adjustment (p=0.04). Comparisons of PRS show that carriers and non-carriers have a similar genetic burden of developing dyslipidemia and CVD, but carriers have lower PRS-based AD risk (p=0.02).

**Conclusions and Relevance:** This study demonstrates that APOECh carriers may have lower levels of apoB and a lower risk of AD. Cohorts with enriched cases are needed to further investigate whether the protective effect is linked to APOE genotypes or other factors.

**Key Points:** *Question:* How does APOE Christchurch (APOECh) variant influence risk of Alzheimer’s disease (AD), cardiovascular disease, and dyslipidemia?

*Findings:* APOECh carriers do not have an elevated risk of dyslipidemia. However, carriers have lower levels of apolipoprotein B (apoB). Comparing polygenic risk scores of carriers vs. noncarriers of APOECh shows carriers are at a lower risk of developing AD before but not after adjusting for multiple comparisons.

*Meaning:* Carriers of APOECh may have a lower risk of developing AD due to lower apoB; however, further epidemiological and model organism studies are needed to validate these observations from the current study.

## Introduction

The *APOE* gene is a known genetic risk factor for neurodegeneration and cardiovascular disease (CVD). APOE is commonly characterized by 3 alleles (ε2, ε3, ε4) and 6 genotypes (ε2/ε2, ε2/ε3, ε2/ε4, ε3/ε3, ε3/ε4, ε4/ε4). The 3 alleles have a population-based frequency of 8.4%, 77.9%, and 13.7%, respectively ^1^. The ε3 allele is the most common allele and does not contribute to disease risk. The ε2 allele binds poorly to the low-density lipoprotein (LDL) receptor and elevates atherogenic lipoprotein levels, whereas the ε4 allele elevates LDL levels by preferentially binding to triglyceride-rich, very low-density lipoproteins (VLDL) ^2^. The ε2 allele has been previously reported to be protective for CVD ^3^ and a majority of APOE ε2 carriers have normal or hypolipemic profile ^4,5^. About 10-15% of individuals with APOE ε2 homozygosity have type III hyperlipoproteinemia (HLP) ^2,6-9^, a condition also associated with *APOE* mutations and deficiency. Several studies reported that ε2 allele significantly increases the risk of hemorrhage in the presence of cerebral amyloid angiopathy ^10-12^. APOE ε2 allele is considered neuroprotective ^13^, in both early and later stages of Alzheimer’s disease (AD) ^14,15^. It is also well-established that ε4 causes neurodegeneration outcomes ^16,17^, where disruption in lipid metabolism may contribute to disease risk ^18^.

Beyond the known effects of APOE ε2 and APOE ε4, several rare and protective APOE variants have been identified recently ^19,20^, including the R154S Christchurch (APOECh) variant in the receptor binding domain and V236E Jacksonville and R251G mutations located in the C-terminal domain. The ultra-rare APOECh mutation (NM_000041.4(APOE):c.460C>A (p.Arg154Ser)), also referred to as p.Arg136Ser or R136S in previous publications ^21,22^, has been hypothesized to protect from *PSEN1*-based autosomal dominant Alzheimer’s disease (AD) ^21^. The protective effect was observed for a single homozygous individual but not for heterozygous descendants ^21^. Functional studies revealed that the R154S variant reduced binding to lipoprotein receptors and heparan sulfate proteoglycans (HSPGs), extracellular matrix proteins that have been suggested to promote amyloid beta (Aβ) aggregation and neuronal uptake of extracellular tau, as well as reduced Aβ42 aggregation *in vitro* ^21^. The V236E protective variant was reported in two studies ^23,24^ to reduce apoE aggregation and to enhance APOE lipidation in human brain as well as reduce amyloid pathology, Aβ plaque-related immune response, and neuritic dystrophy in murine models. The function of the newly identified R251G variant ^25^ remains to be studied.

It is important to understand whether other phenotypic manifestations can be caused by those protective variants. For example, mutations in the LDL-receptor binding domain of the *APOE* gene, including the APOECh mutation, have been associated with type III HLP ^2,6-9^, and premature cardiovascular disease (CVD) ^26^. The APOECh mutation was first discovered in a man with APOE ε2/ε2 genotype and type III HLP ^9^. Preclinical studies using animal models and human cellular models are needed to address specific effects of APOE variants in lipid biology in general, and the particular impact of APOECh on neuropathological features including amyloid plaques and tau tangles, immune response, vascular integrity and function, and other AD-related pathways ^19,20^. To date, few studies have reported on the effect of APOECh on AD and other phenotypic traits^21,22,27^. This study examined the UK Biobank (UKB) whole exome sequencing (WES; N=454,756) and whole genome sequencing (WGS; N=141,948) data to characterize the phenotypic spectrum of APOECh carriers.

## Methods

### Participants

The UKB is a prospective cohort study with deep genetic and rich phenotypic data on nearly half a million individuals ^28^. At the time this study was completed, WES and WGS data were available for ∼450,000 ^29^ and ∼150,000 individuals, respectively ^30^. The UKB study was conducted under generic approval from the NHS National Research Ethics Service (approval letter dated 17th June 2011, Ref 11/NW/0382). All participants gave full informed written consent. Details on the data release version and sequencing are described in **eMethods in Supplement**.

### Genotype Screening

*APOE* genotype (**eTable 1 in Supplement**), *APOE* protective variants, and mutations for neurodegenerative disorders with evidence of Mendelian inheritance in *APP, PSEN1, PSEN2, GRN, SORT1, MAPT*, and *APBB2* were screened for both carriers and noncarriers of APOECh using WES and WGS data (**eTable 2 in Supplement**). In *APOE*, we examined rs429358, rs7412, and the missense APOECh variant rs121918393-A. We also examined whether any of the APOECh carriers had V236E Jacksonville (rs199768005-A) and R251G (rs267606661-G) protective alleles.

### Study Design and Statistical Analysis

In this study (**Figure 1**), we compared the clinical profiles of 36 European APOECh carriers and 129,240 matched noncarriers for 11 binary traits, 80 quantitative traits, and 10 polygenic risk scores (PRS) described in Phenotype Definitions (**eMethods in Supplement**). To avoid confounding by genetic ancestry and due to a lack of available clinical information, the only APOECh carrier with admixed American ancestry was excluded from downstream statistical analyses. We identified a noncarrier cohort by maximizing the number of matching sets that could be generated from among UKB participants given age (5-year bin), genetic sex, and genetic ancestry from Pan-UKBB (UKB Return Field 31063) ^31^ resulting in a 1:3,590 ratio (36 carriers:129,240 noncarriers).

**Figure 1.**
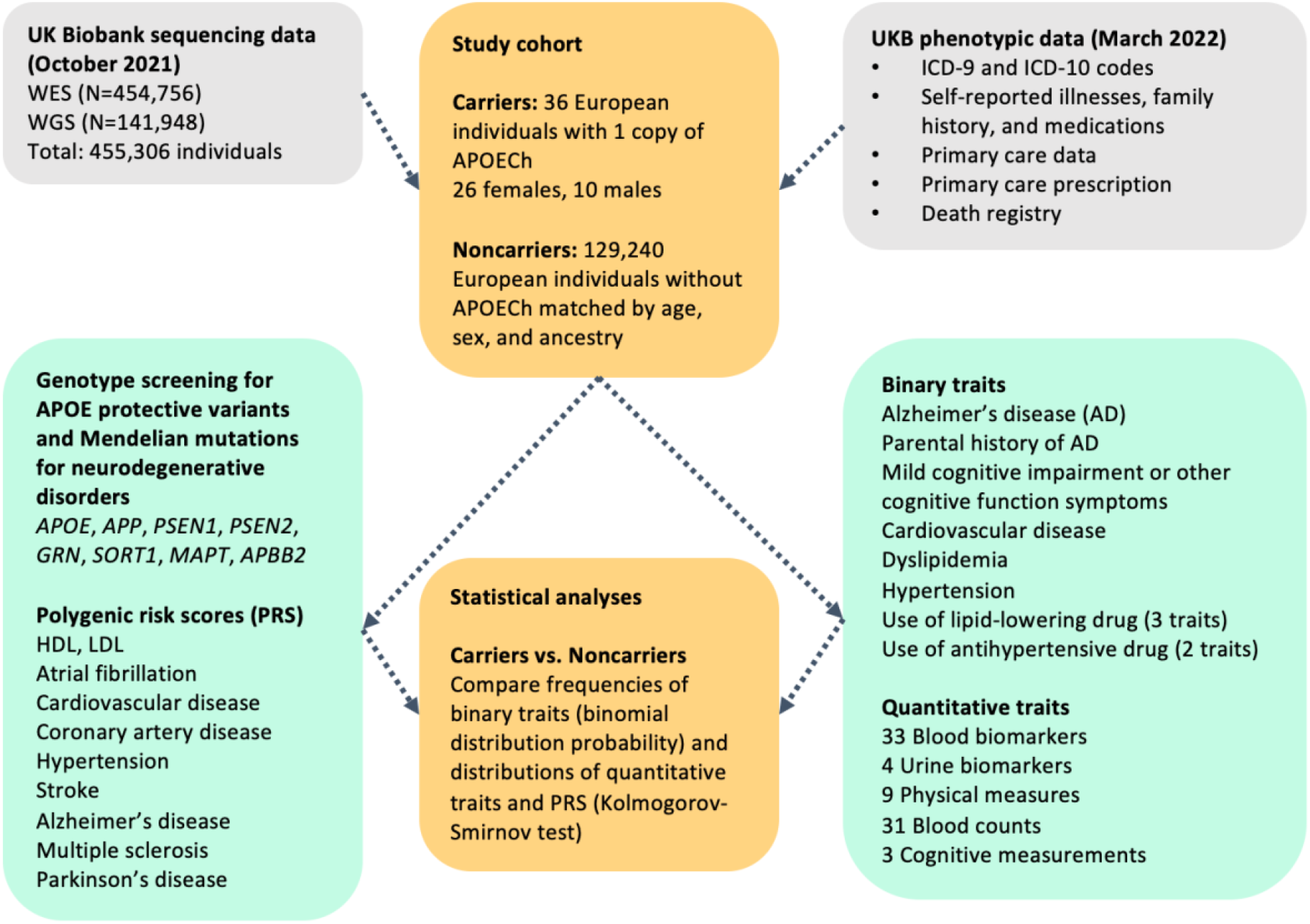
Schematic of study design

Medical history, including ICD-10 diagnoses (UKB Data Field 41270), ICD-9 diagnoses (UKB Data Field 41271), ICD-10 primary cause of death (UKB Data Field 40001), ICD-10 secondary cause of death (UKB Data Field 40002), self-reported non-cancer illness (UKB Data Field 20002; **eTable 3 in Supplement**) and medications (UKB Data Field 20003; **eTable 4 in Supplement**), as well as primary care data and prescriptions (gp_scripts) ascertained from baseline assessment center visit were evaluated to determine whether specific medical characteristics are enriched in the APOECh carriers. Only 9 carriers had metabolomics, 7 had proteomics, and 3 had brain imaging data (**eTable 5 in Supplement**). Given only 37 carriers, of which 36 Europeans are retained for analysis, we focused on assessing the difference between carriers and noncarriers in medical history for AD and related cognitive traits, cardiovascular health, and quantitative traits. Additionally, we explored whether PRS differed between carriers and noncarriers to determine whether their genetic background for developing cardiovascular and neurodegenerative diseases could differentially influence disease risk.

For binary traits assessed in this study, the cumulative probability function was calculated using R to assess whether carrier event frequency is lower or higher than expected based on noncarrier prevalence. For quantitative traits, a two-sided Kolmogorov-Smirnov (KS) test in R (ks.test) was used to test whether the distributions for carriers vs. noncarriers differed significantly. Baseline values measured at enrollment were used for quantitative traits as they have the greatest number of observations. Longitudinal effects were not assessed due to missingness and loss to follow-up.

### Phenotype Definitions

We evaluated 11 binary traits, including AD, family history of AD, mild cognitive impairment (MCI) or other cognitive function symptoms, dyslipidemia, use of lipid-lower drugs, self-reported statin, hypertension, antihypertensive use, and CVD. We also assessed 80 quantitative traits, including blood and urine-based biomarkers. Details are provided in **eMethods in Supplement**.

## Results

We identified 36 heterozygous individuals by WES and 7 by WGS, of whom 6 were sequenced by both, resulting in APOECh allele frequency of 4.06×10^−5^. To date, no individuals homozygous for the APOECh have been observed in the UKB sequencing data. Among APOECh carriers, 36 are of European ancestry and 1 individual is of admixed American ancestry based on genetic ancestry assessed by Pan-UKBB (UKB Return Field 31063) ^31^. These observations are consistent with the allele frequency observed in the gnomAD database. For example, gnomAD ^32^ v2.1.1 includes 2 APOECh heterozygous individuals in 77,963 WES samples (allele frequency [AF] = 1.28×10^−5^). The gnomAD ^32^ v3.1 database includes 7 APOECh heterozygous individuals in 76,065 WGS samples (AF = 4.6×10^−5^), of whom 3 have Non-Finnish European ancestry and 4 have Latino/admixed American ancestry.

The 37 APOECh carriers are comprised of 27 females and 10 males. The demographic characteristics of 37 carriers of the APOECh variant are described in **Table 1**. The carriers had a median age of 68 as of January 1, 2022, or at death (UKB Data Field 40007). Upon examining clinical history, self-reported noncancer illnesses and parental medical history (see **Methods** and **eMethods in Supplement** for codes and UKB data field IDs) in 36 European carriers, none of these individuals had any form of dementia or mild cognitive impairment (not significantly different from noncarriers, p=0.83 and p=0.81, respectively; **eTable 7 in Supplement**). Two individuals have paternal history of AD, and two others have maternal history of AD (not significantly different from noncarriers, p=0.74; **eTable 7 in Supplement**). There was no difference between carriers and noncarrier in FIS, RT, or EDU_YR, as proxy phenotypes for cognitive function (**eTable 7 in Supplement**). The admixed American APOECh carrier does not have any hospital records for the variables assessed in this study. Clinical characteristics are not provided to protect the identity of this individual.

**Table 1.** Characteristics of APOE Christchurch variant carriers and matched noncarriers in UKB European samples

|  | Carriers (N=37) | European Noncarriers (matched to 36 European carriers with 1:3590 ratio; N=129,240) |
| --- | --- | --- |
| *Age* | Median: 68.63<br>Mean: 68.91<br>Min: 56.62<br>Max: 82.06 | Median: 68.38<br>Mean: 68.86<br>Min: 56<br>Max: 82.97 |
| *Genetic sex* | 27 females<br>10 males | 93,340 females<br>35,900 males |
| *Pan-UKBB derived ancestry* | 36 Europeans<br>1 admixed American | 129,240 Europeans |
| *Deceased participants* | 2 deceased<br>Primary causes of death: acute myocardial infarction (MI), hypertensive heart disease without heart failure | 8,779 deceased<br>Primary causes of death: lung cancer (9.4%), breast cancer (5.8%), pancreatic cancer (3.8%), acute MI (3.3%), chronic ischaemic heart disease (2.7%) |
| *Alzheimer's disease (ICD + self-report)* | None | 660 (0.51%)<br>643 ICD code; 39 self-reported |
| *Parental history of Alzheimer's disease* | 4 (10.8%) with at least 1 parent (2 paternal; 2 maternal) | 17,428 (13.5%) with at least 1 parent (6500 paternal; 11705 maternal) |
| *Mild cognitive impairment or other cognitive symptoms ICD codes* | None | 755 (0.58%) |

It is important to consider the effect of the APOECh in the context of other AD risk or protective variants. Five carriers are heterozygous for APOE ε4, 31 carriers are homozygous for APOE ε3, and one carrier has the ε2/ε3 genotype (**eTable 1 in Supplement**). We find that the APOECh carriers are enriched for the ε3/ε3 genotype. Like APOE ε2, APOECh has been reported to reduce the aggregation of amyloid plaques and has been reported to have a protective effect due to loss of *APOE* function ^21^. Thus, the APOECh carrier with APOE ε2/ε3 genotype may have greater protective effect against AD.

We screened for *APOE* protective variants as well as mutations with evidence of Mendelian inheritance in *APP, PSEN1, PSEN2, GRN, SORT1, MAPT*, and *APBB2* to assess the genetic risks of carriers and noncarriers. We identified APOECh carriers with *GRN* rs5848-T, *SORT1* rs12740374-T, and *APBB2* rs13133980-G (**eTable 2 in Supplement**). However, these variants have low penetrance or mixed evidence as AD risk loci. No APOECh carriers had the protective V236E Jacksonville and R251G variants (**eTable 2 in Supplement**). We found that the carriers had a lower overall PRS for AD (p=0.02), but not for Parkinson’s disease (p=0.06) or multiple sclerosis (p=0.33) (**eTable 7 in Supplement**).

Given the important role of *APOE* and the hypothesized role of APOECh in dyslipidemias and CVD risk, we examined the cardiovascular and metabolic health of APOECh carriers. One participant died at the age of 72 years from acute myocardial infarction (primary cause of death, UKB Data Field 40001) and atherosclerotic heart disease of native coronary artery (secondary cause of death, UKB Data Field 40002). Another participant died at the age of 67 from hypertensive heart disease (primary cause of death, UKB Data Field 40001). However, there is no difference in occurrence of CVD (p=0.34), dyslipidemia (p=0.13) and hypertension (p=0.24) as defined based on ICD codes and self-reported data when comparing all carriers to noncarriers. Our analyses demonstrated that the polygenic risk scores for HDL, LDL, and polygenic risk for atrial fibrillation, cardiovascular disease, coronary artery disease, hypertension, and stroke do not differ between carriers and noncarriers (**eTable 7 in Supplement**). Furthermore, linear and logistic regression analyses with polygenic score (HDL and LDL) or polygenic risk score (CVD, hypertension, and AD) as a covariate for quantitative and binary traits, respectively, showed that there is no difference in HDL/LDL levels, CVD, and AD between carriers and noncarriers (**eTable 8 in Supplement**).

Additionally, there is no statistically significant difference in self-report of statin use (p=0.46), prescribed lipid-lowering drugs (p=0.30), and self-reported/prescribed lipid-lowering drugs (p=0.34), as well as use of antihypertensives based on self-report only (p=0.15) and self-reported/prescribed antihypertensives (p=0.29; **eTable 7 in Supplement**). As a sensitivity analysis to address the limitation of primary care prescription data being available in half of the UKB cohort, we examined whether there is a difference in medication use in a subset of carriers and matched controls who have primary care prescription data (**eMethods in Supplement**). We found no difference in the use of lipid-lowering and antihypertensive medications (**eTable 4 in Supplement**).

There is no significant difference between carriers and noncarriers for most blood-based lipid biomarkers measured at baseline (**Table 2, eTable 9 and eFigure 1 in Supplement**), both adjusted and unadjusted for self-reported statin use at the time of recruitment, except for apoB (statin unadjusted p=0.004, statin adjusted p=0.036). There were no significant differences in physical measures and urine biomarkers (**eTable 9 and eFigures 2 and 3 in Supplement**). Although microalbumin levels showed nominal significant difference of p<0.05, 71.4% of individuals assessed have missing or non-reportable data. When microalbumin is treated as a categorical variable defined as < 30mg/L (normal), 30-300mg/L (microalbuminuria), and > 300mg/L (macroalbuminuria), there are no statistical differences between carriers and noncarriers. Among non-lipid blood biomarkers and hematological traits, a few traits showed difference at nominally significant level (p<0.05) but were not significant after accounting for multiple testing correction for 22 non-lipid blood biomarkers or 31 hematological traits (**eTable 9 and eFigures 4 and 5 in Supplement**). Briefly, carriers exhibit higher aspartate aminotransferase (p=0.029), higher erythrocyte distribution width (p=0.038), higher thrombocyte volume (p=0.017), and lower high light scatter reticulocyte count (p=0.03).

**Table 2.** Blood-based lipid biomarkers of APOECh carriers and noncarriers

|  | EUR Carriers<br>(N=36) |  | EUR Noncarriers<br>(N=129,240) |  | KS test |
| --- | --- | --- | --- | --- | --- |
|  | Min, Max | Median | Min, Max | Median |  |
| *Apolipoprotein A* | 1.03-2.12 | 1.64 | 0.52-2.50 | 1.55 | D=0.19 P-value=0.239 |
| *Apolipoprotein B* | 0.65-1.37 | 0.88 | 0.40-1.99 | 1.02 | D=0.31 P-value=0.004 |
| *Apolipoprotein B (statin adjusted)* | 0.65-1.37 |  | 0.40-2.76 |  |  |
|  |  | 0.94 |  | 1.06 | D=0.25 P-value=0.036 |
| *C reactive protein* | 0.09-16.48 | 1.12 | 0.08-78.26 | 1.32 | D=0.12 P-value=0.662 |
| *Cholesterol* | 3.82-7.83 | 5.32 | 1.71-12.50 | 5.73 | D=0.18 P-value=0.263 |
| *Cholesterol (statin adjusted)* | 3.82-7.83 | 5.80 | 1.88-15.56 | 5.91 | D=0.15 P-value=0.427 |
| *High-density lipoprotein* | 0.80-2.05 | 1.56 | 0.23-4.19 | 1.47 | D=0.2 P-value=0.225 |
| *Low-density lipoprotein* | 2.19-5.06 | 3.25 | 0.75-8.86 | 3.55 | D=0.23 P-value=0.072 |
| *Low-density lipoprotein (statin adjusted)* | 2.23-5.06 |  | 0.80-12.96 |  |  |
|  |  | 3.36 |  | 3.70 | D=0.21 P-value=0.119 |
| *Lipoprotein A* | 4.10-178.95 | 22.00 | 3.80-189.00 | 20.50 | D=0.11 P-value=0.838 |
| *Triglycerides* | 0.79-6.31 | 1.71 | 0.23-11.15 | 1.42 | D=0.16 P-value=0.329 |

## Discussion

A curious case study of a woman with the autosomal dominant *PSEN1* E280A mutation with two copies of the APOECh who was protected from developing MCI until her seventies despite unusually high positron emission tomography (PET) measurements of Aβ plaque burden, has nominated the Christchurch variant as a protective mutation ^21^ and led to a question whether mimicking the functional consequence of this mutation pharmacologically or genetically could be a useful tool to further understand the role of *APOE* in neurodegeneration. Investigators from the original case report recently described their *in vivo* follow-up from PET imaging and postmortem findings and identified atypical tau pathology and unusual regional distribution of tau aggregates ^33^. Their findings suggest that APOECh modifies the effect of tau on AD severity, progression, and clinical representation in a regional-specific manner and its protective role is associated with expression in astrocytes and microglia ^33^.

In a previous study, heterozygous carriers of APOECh variant did not show protective effect against AD ^22^; however, given the rarity of the APOECh variant, it has been challenging to investigate the effect of APOECh dosage on AD and other phenotypic traits. Homozygous carriers were not observed in the UKB. Among the heterozygous carriers (age range: 56.62-82.06, median: 68.63), none have developed AD or MCI, including 4 individuals with parental history of AD. While this may suggest that the APOECh carriers are protected, continued follow-up as the cohort ages is necessary as the proportion of noncarriers with AD or MCI indicates that most participants may not be old enough to have developed AD. Additionally, the interaction effect between age and APOECh variant was not able to be assessed. Interestingly, the carriers showed a decreased genetic risk measured by PRS for Alzheimer’s disease (p=0.02), which cannot be attributed to APOECh alone since this rare variant is not included in AD GWAS and polygenic risk score calculations. While only one carrier had one copy of the protective APOE ε2 allele, none of them had the recently described protective V236E Jacksonville and R251G variants (**eTable 2 in Supplement**), and a few were found to carry potential risk alleles in *GRN* and *SORT1*. Meanwhile, it is important to note that 23 noncarriers had 1 copy of APOE V236E and 17 noncarriers had 1 copy of APOE R251G, both of which were recently shown to be protective against AD ^25^. Larger future studies such as All of US (N∼1 million) or Our Future Health (N∼5 million) may be able to assess LD and/or an interaction effect of the APOECh variant with other AD-protective variants resulting in heterozygous carriers having lower AD polygenic risk.

Previous reports have shown both APOE ε2 homozygosity and APOE3-Christchurch are associated with type III HLP ^6,9^, a risk factor for atherosclerosis, that is characterized by elevated cholesterol, triglycerides, and VLDL. However, we did not observe differences in lipid biomarkers among carriers vs. noncarriers in this study, whether unadjusted or adjusted for statin use.

Among the lipid biomarkers assessed in this study, both statin-adjusted and unadjusted apoB levels were significantly lower in carriers compared to noncarriers (**Table 2**). While statin adjustment slightly attenuated the difference in apoB levels, the significant difference persisted, suggesting that lower apoB may be a characteristic of APOECh carriers. A study reported that higher levels of plasma apoB are associated with cognitive decline ^34^ and a recent study has demonstrated that cerebrospinal fluid apoB levels are correlated with tau pathology in pre-symptomatic individuals and elevated in AD patients ^35^. Together, this suggests that having lower apoB may be protective for AD.

## Limitations

This study includes several limitations. We did not identify any homozygotes for APOECh and are unable to validate the findings reported by Arboleda-Velasquez and colleagues ^21^. Due to the rarity of the APOECh variant, the sample size of carriers is insufficient for leveraging UKB brain imaging, metabolomics, and proteomics data, which are currently only available on partially overlapping subsets of UKB participants; the overlap with carriers was too small to perform statistical analyses in this study (**eTable 5 in Supplement**). For the quantitative traits assessed, only baseline measurements were considered given that repeated biomarkers measurements are available only for a subset of UKB participants. Statin adjustment for lipid biomarkers were only applied using self-reported statin at baseline. Primary care prescription data were not considered for statin adjustment because only 48.8% of the UKB cohort has prescription data (**eTable 9 in Supplement**). Statin adjustment may be incomplete because not all individuals report medication use; confounders assessed by self-report may be sub-optimally controlled depending on the outcome of interest ^36^.

## Conclusions

We have developed a framework to profile APOECh carriers, and this approach can be applied to assess other rare variants. To our knowledge, this work has examined the clinical phenotypes in the largest cohort of APOECh carriers among UKB participants. We found lower levels of apoB in APOECh carriers which could partially be due to lipid-lowering medication use, but the difference is not fully attenuated by lipid-lowering medication adjustment; however, potential limitations about self-report accuracy could impact this finding. We also find more individuals on lipid-lowering medications, suggesting that APOECh carriers may have higher lipid levels, which would be consistent with previous case reports. While the APOECh variant is very rare and larger cohorts are needed to assess its contribution to dementia, dyslipidemia, and CVD, the UKB provides an unprecedented opportunity to follow these carriers and elucidate the underlying role of APOECh in disease etiology.

## Data Availability

All data produced in the present work are contained in the manuscript and/or supplement. Data used in the preparation of this article were obtained from the UK Biobank (https://www.ukbiobank.ac.uk/researchers/). All bona fide researchers from academic, commercial, and charitable organizations can apply access to the UK Biobank resource to conduct health-related research that is in the public interest.

## Notes

### Competing Interest Statement

All authors are employees at the Janssen Pharmaceutical Companies of Johnson & Johnson at the time this work was performed. Simon Lovestone is a founding director of Akrivia Health Ltd (UK). Mary Helen Black is currently an employee at Foresite Labs.

### Funding Statement

This study did not receive any funding.

### Author Declarations

The UK Biobank study was conducted under generic approval from the NHS National Research Ethics Service (approval letter dated 17th June 2011, Ref 11/NW/0382). All participants gave full informed written consent.

