## Supplemental Methods for "Characterization of *APOE* Christchurch carriers in 455,306 UK Biobank participants"

### *Participants*

This research has been conducted using the UK Biobank—project number 52293. Phenotypic data were derived based on the ukb50945 March 2022 data release, while education years (EDU_YR), fluid intelligence score (FIS), and reaction time (RT) were extracted from ukb49194 October 2021.

### *Sequencing*

WES data for UK Biobank participants were generated at the Regeneron Genetics Center (RGC) as part of a public-private partnership between eight pharmaceutical companies (AbbVie, Alnylam Pharmaceuticals, AstraZeneca, Biogen, Bristol-Myers Squibb, Pfizer, Regeneron, and Takeda) and UK Biobank. Briefly, genomic DNA underwent paired-end 75bp WES at RGC using the IDT xGen v1 capture kit on NovaSeq6000 machines. Initial QC was performed by RGC and included checks for sex discordance, contamination, unresolved duplicate sequences, and discordance with microarray genotyping data. Detailed methods for joint variant calling, QC thresholds, and filtering criteria have been previously described [1]. WES data from 450,000 UK Biobank participants were made publicly available in October 2021 and underwent internal QC and filtration in a manner similar to that previously described prior to ingestion and analysis. WGS data for UK Biobank participants were generated from sequencing efforts at the Wellcome Sanger Institute and deCODE as part of a public-private partnership involving Amgen, AstraZeneca, GlaxoSmithKline, and Janssen Pharmaceutical Companies of Johnson & Johnson, alongside Wellcome and UK Research and Innovation (UKRI). Briefly, 150 bp paired-end sequencing was performed on an Illumina NovaSeq 6000. Per-sample quality checks were performed (e.g., yield, read count, GC fraction, insert fragment size distribution, etc.), with samples passing checks proceeding to genotyping. Data were aligned to the GRCh38 reference genome. Samples were checked for contamination, sex mismatch, and concordance with microarray-called genotypes. Joint-genotyped variants were called using the Genome Analysis Toolkit (GATK) and Graphtyper; we focus on the GATK-called dataset here. Data underwent a multistep QC process. For autosomes, hemizygous genotypes were masked, as were any genotypes with genotype qualities (GQ) less than 30. Indel genotype calls were masked if they had a depth (DP) less than 10; SNP calls were masked if they had a DP less than 20. Additionally, variants (sites) were subjected to a series of filters following the Broad Institute's recommendations for hard filtering (SNPs: QD < 2, QUAL < 30, SOR > 3, FS > 60, MQ < 40, MQRankSum < -12.5, ReadPosRankSum < -8; indels: as before, but with FS > 200 and ReadPosRankSum < -20; per-site missingness > 10%). The same filters were applied for the X chromosome, but variant genotypes were masked if they had a DP less than 10. WGS data from 141,948 UK Biobank participants were made available to us in July 2021 after internal QC (including removal of duplicated variants).

***Phenotype Definitions***

The phenotype data collected includes self-reported information, standardized assessments, hospital and primary care records, cancer and death registries, quantitative biomarkers, infectious disease antigens, and multi-modal imaging results for up to 500,000 individuals. Birth date was derived using year and month of birth (UKB Data Field 34 and 52). Current age is based on the individuals' age as of January 1, 2022; age-at-death was assessed for deceased subjects using UKB Data Field 40007.

AD is defined as ICD-10 codes F00, F00.1, F00.2, F00.9, G30, G30.0, G30.1, G30.8, and/or G30.9 in UKB Data Fields 41270 (ICD-10 diagnoses), 40001 (ICD-10 primary cause of death), and 40002 (ICD-10 secondary cause of death), ICD-9 code 331 in UKB Data Field 41271 (ICD-9 diagnoses), and self-reported code 1263 in UKB Data Field 20002 (self-reported non-cancer illness). Family history (FH) of AD was ascertained using self-reported code 10 from UKB Data Fields 20110 and 20107 (illnesses of mother and father, respectively); the subject is characterized as having FH if at least 1 parent was reported to have AD. MCI and other cognitive function symptoms were defined using ICD-10 codes F06.7, R41, and R41.8 and ICD-9 code 3101 in UKB Data Fields 41270, 40001 40002, and 41271. CVD was broadly defined as any ICD-10 codes in chapter 9 I00-I99 (diseases of the circulatory system), chapter 18 R00-R09 (symptoms and signs involving circulatory and respiratory systems), or chapter 21 Z867 (personal history of disease of the circulatory system), ICD-9 codes in chapter 7 (390-459), chapter 16 785-786 (symptoms involving cardiovascular system, respiratory system, and other chest symptoms), and self-reported history of any cardiovascular conditions (UKB Data Field 20002; **eTable 3 in Supplement**). Dyslipidemia was defined as ICD-10 codes E78 and E78.0-E78.9, ICD-9 codes 2720-2729, and self-reported code 1473 for high cholesterol (UKB Data Field 20002). Use of lipid-lowering drugs include primary care prescription data from any time point under ATC codes beginning with prefix C10 and any self-reported use of statins and other types of non-statin lipid-lowering drugs (UKB Data Field 20003; **eTable 4 in Supplement**). Hypertension was defined as ICD-10 code I10 (essential hypertension) and ICD-9 codes for 401, 4010, 4011, and 4019 (essential hypertension). Use of antihypertensive include primary care prescription data from any time point under ATC codes beginning with prefixes C02, C03, C07,

C08, and C09 and any self-reported use of antihypertensive drugs (UKB Data Field 20003; **eTable 4 in Supplement**).

A total of 80 quantitative traits were assessed (**eTable 6 in Supplement**). Blood and urine biomarkers, hematologic traits, physical measures, education years, and cognitive traits (fluid intelligence score, and reaction time) were ascertained at assessment centers and the first instance at baseline were used for evaluation. LDL, cholesterol, and apoB have been adjusted for self-reported statin at baseline as described in the study published by Sinnott-Armstrong and Tanigawa [2]. Systolic and diastolic blood pressure have been adjusted for self-reported antihypertensive at baseline [3]. Primary care prescription use of other types of lipid-lowering drugs and antihypertensive were not considered for medication adjustment given primary care is only available for approximately half of the entire UKB cohort. Quantitative traits with multiple measurements from the same visit (e.g., blood pressure) were averaged before comparing carriers to noncarriers. Education attainment was converted to education years based on UKB Data Fields 6138 (educational qualifications) and 10722 (education qualifications pilot); the maximum value is considered as the highest education completed [4]. Additionally, standard polygenic risk scores [5] for 2 lipid traits (high-density lipoprotein [HDL] and low-density lipoprotein), 5 cardiovascular events (atrial fibrillation, cardiovascular disease, coronary artery disease, hypertension, and ischaemic stroke), and 3 neurological diseases (Alzheimer's disease, multiple sclerosis, and Parkinson's disease) were extracted from ukb669060 (ascertained August 2022) and compared to assess whether genetic risk differs between carriers and noncarriers.

***Medical Records Data Availability***

Phenotypic data availability varies across individuals in the UKB. Thirty out of thirty-seven APOECh carriers have at least 1 ICD-9 (UKB Data Field 41271) or ICD-10 code (UKB Data Field 41270), 24 have at least one self-reported non-cancer illness at the time of baseline assessment (UKB Data Field 20002), 30 have at least one self-reported medication at the time of baseline assessment (UKB Data Field 20003), 18 have at least one primary care record and primary care prescription record (gp_scripts), and 32 have at least 1 lipid biomarker measurement at baseline (UKB Data Fields 30630, 30640, 30710, 30690, 30760, 30780, 30790, 30870). The proportion of data available for each category is shown in **eTable 10 in Supplement**.

***Sensitivity analysis of medication usage in individuals with prescription data***

Given primary care prescription data is only available for 48.8% of the UKB cohort, we performed a sensitivity analysis to assess whether there is a difference in medication use among carriers and noncarriers with prescription data. We further subsetted the study sample cohort to 17 European APOECh carriers and 27,013 matched noncarriers with prescription data. The 27,013 noncarriers included for assessment were extracted from the pool of 129,240 noncarriers that were originally matched based on sex, age, and ancestry presented as the primary analysis of this study. We compared the frequencies of lipid-lowering and antihypertensive drugs in carriers and non-carriers using binomial distribution probability.
