## Supplemental Tables and Figures for "Characterization of *APOE* Christchurch carriers in 455,306 UK Biobank participants"

#### Supplemental Figures

eFigure 1. Cumulative distributions of lipid biomarkers and Kolmogorov-Smirnov test results

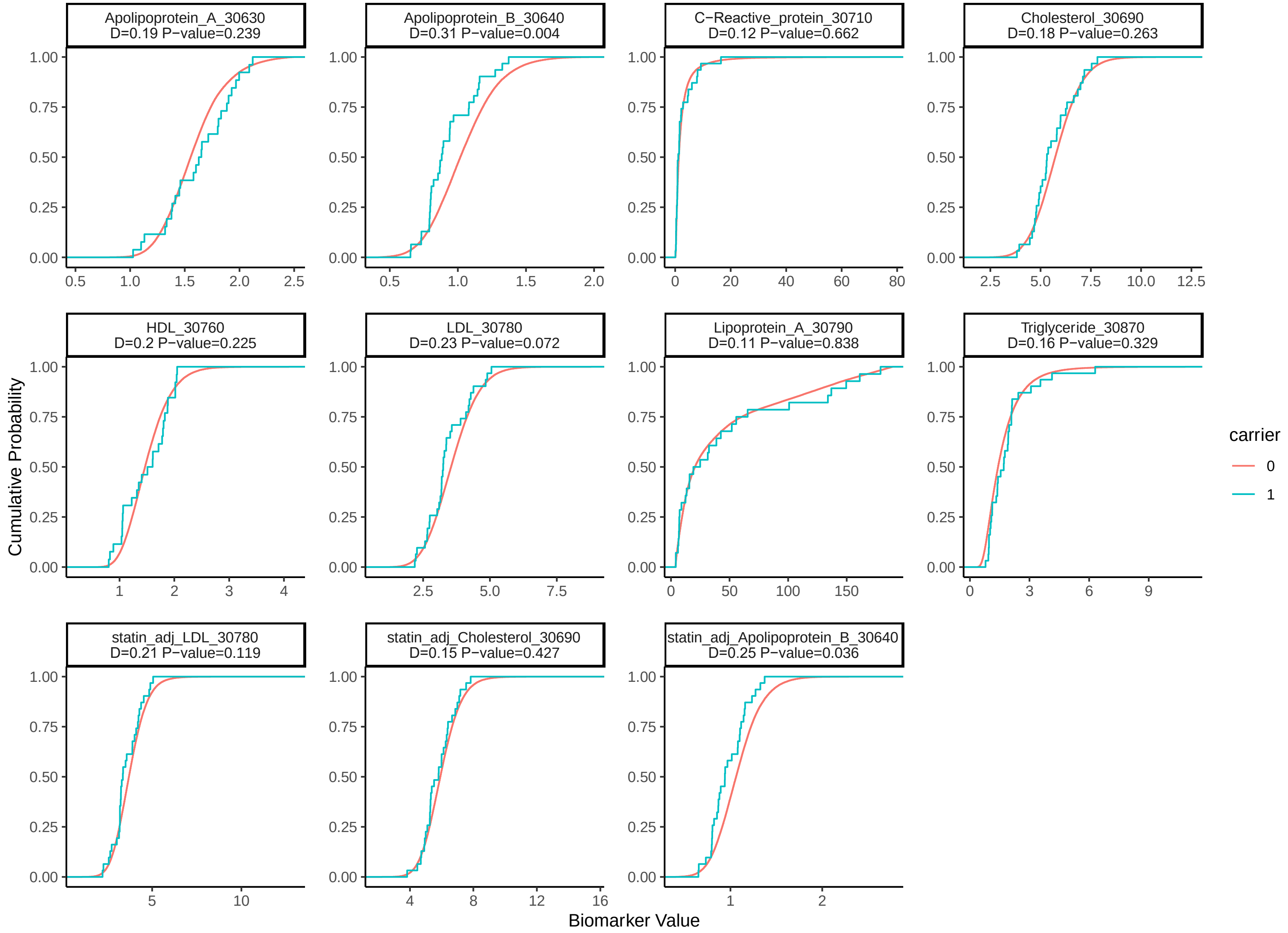

eFigure 2. Cumulative distributions of physical measures and Kolmogorov-Smirnov test results

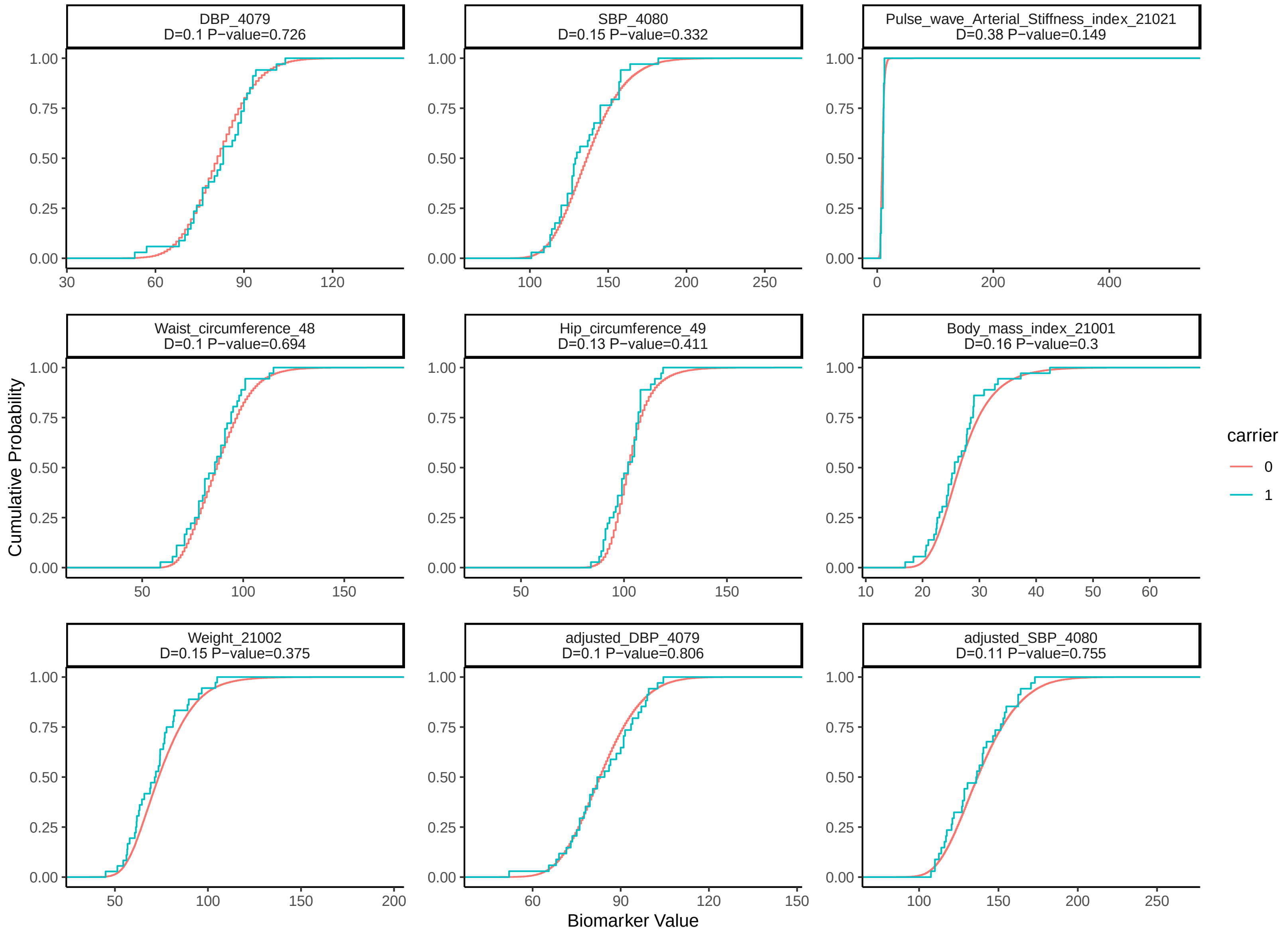

eFigure 3. Cumulative distributions of urine biomarkers and Kolmogorov-Smirnov test results

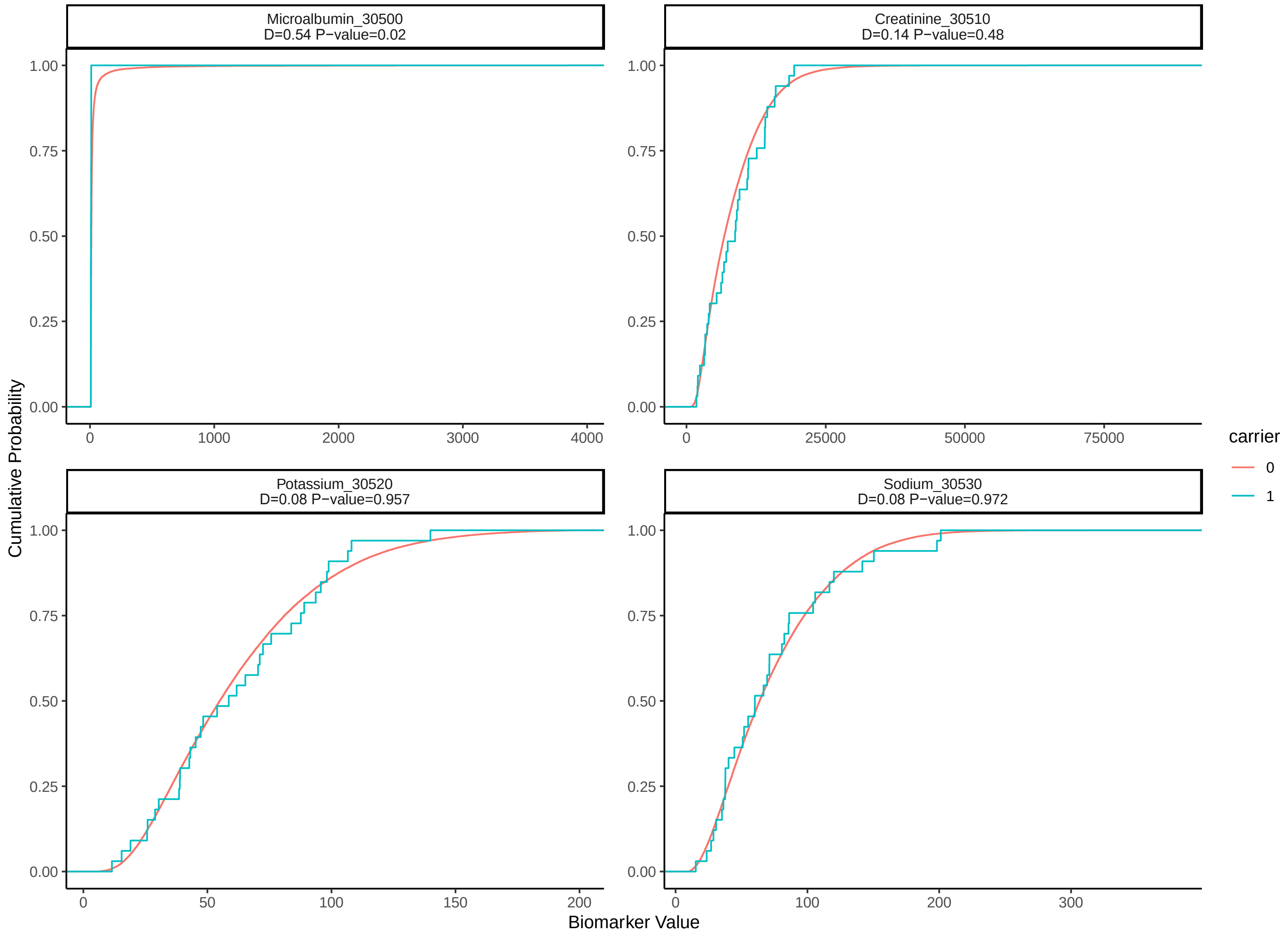

eFigure 4. Cumulative distributions of blood biomarkers and Kolmogorov-Smirnov test results

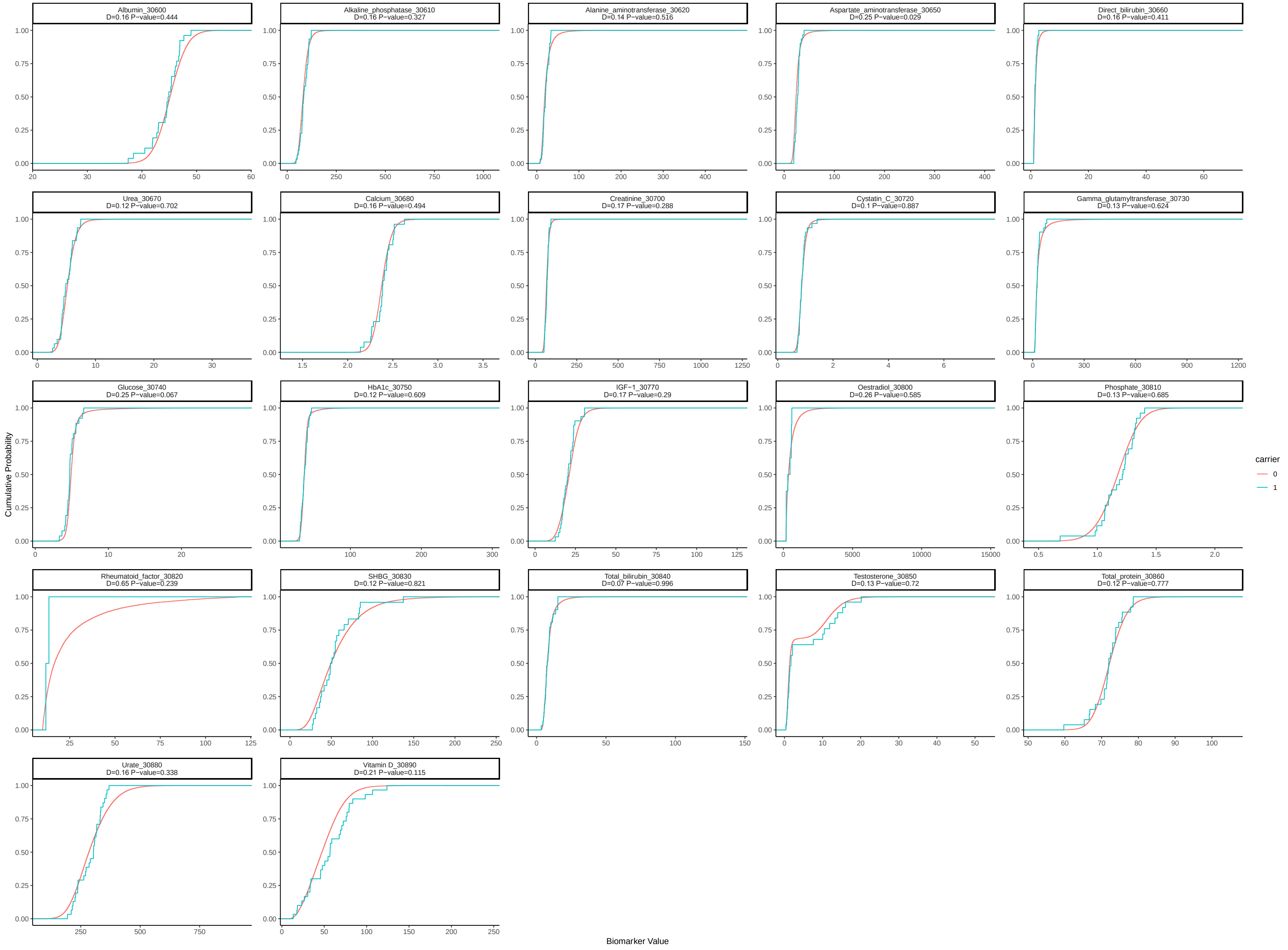

### eFigure 5. Cumulative distributions of hematological traits and Kolmogorov-Smirnov test result

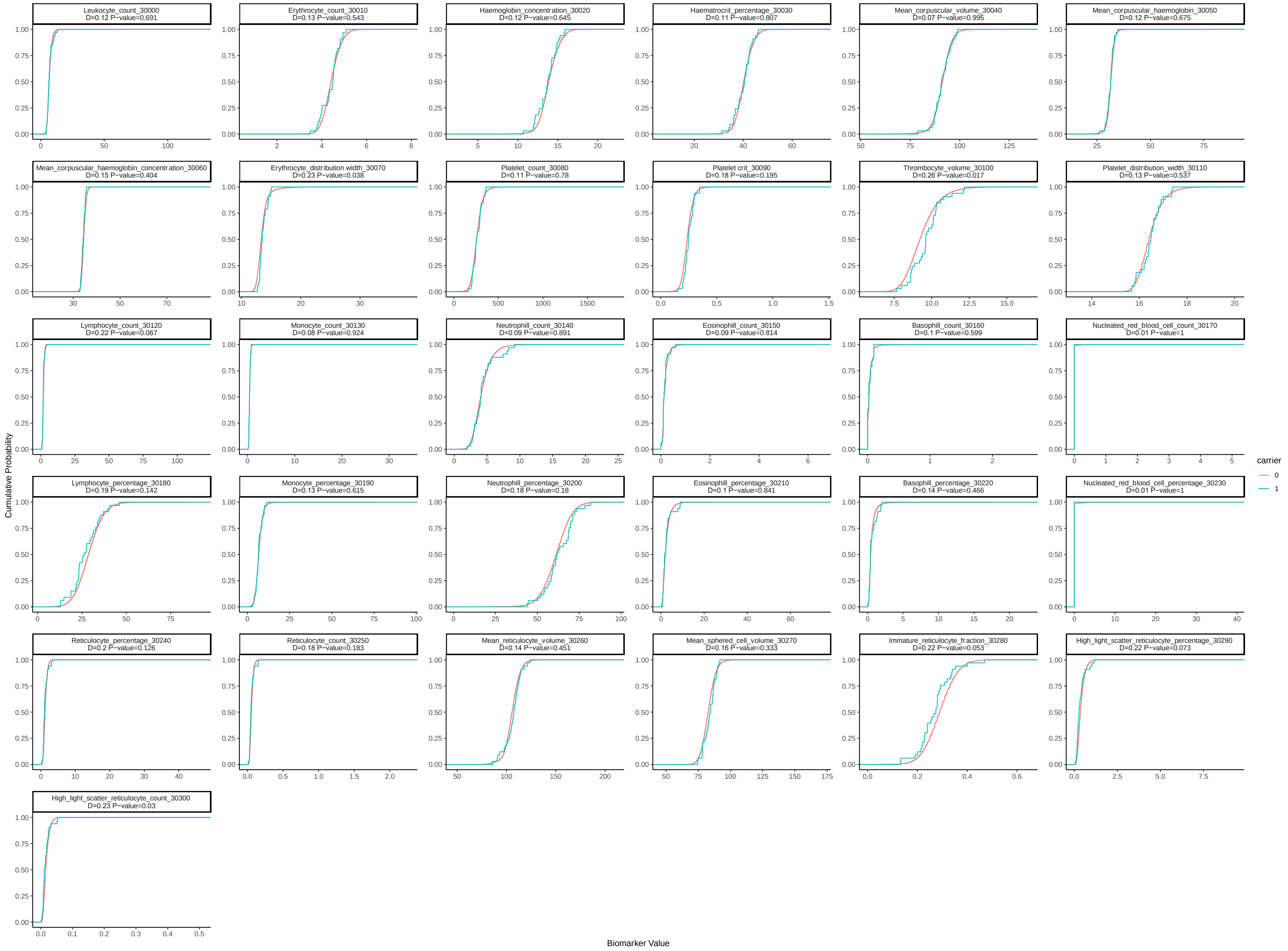

#### Supplemental Tables

eTable 1. *APOE* genotype of APOECh carriers and matched noncarriers

| *Genotype* | *rs429358* | *rs7412* | *APOECh carriers (N=36)* | *Noncarriers (N=129,240)* | *Noncarriers with AD (N=660)* | *Probability of genotype enrichment in carriers* |
|---|---|---|---|---|---|---|
| *e2/e2* | T/T | T/T | 0 | 518 (0.4%) | 1 (0.2%) | 0.87 |
| *e2/e3* | T/T | C/T | 1 (2.8%) | 15,887 (12.3%) | 28 (4.2%) | 0.99 |
| *e2/e4 or e1/e3* | C/T | C/T | 0 | 3,216 (2.5%) | 18 (2.7%) | 0.40 |
| *e3/e3* | T/T | C/C | 30 (83.3%) | 75,200 (58.2%) | 209 (31.7%) | 0.001 |
| *e3/e4* | C/T | C/C | 5 (13.9%) | 30,781 (23.8%) | 302 (45.8%) | 0.95 |
| *e4/e4* | C/C | C/C | 0 | 2,785 (2.2%) | 92 (13.9%) | 0.46 |
| *NA* | Missing | | 0 | 853 (0.7%) | 10 (1.5%) | 0.79 |

eTable 2. Screening APOECh carriers for APOE protective variants and mutations reported for neurodegenerative disorders with a Mendelian inheritance

| *Gene* | *SNPs assessed* | *SNPs found in UKB data* | *APOECh carriers* | *APOECh noncarriers* | *Additional annotations* |
|---|---|---|---|---|---|
| *APOE* | rs199768005<br>rs267606661 | rs199768005<br>rs267606661 | None | rs199768005: 23 individuals with 1 copy<br>rs267606661: 17 individuals with 1 copy<br>No individuals with both variants | *APOE* rs199768005-A is also known as V236E Jacksonville variant<br>rs267606661-G is also known as R251G<br>Both rare variants are protective for AD [1]. |
| *APP* | rs63750264<br>rs63749964<br>rs63750671<br>rs63750066<br>rs63750399<br>rs63750734<br>rs63751039<br>rs63750973<br>rs63750643<br>rs193922916<br>rs63750847 | rs63750264<br>rs63750066<br>rs63750847 | None | rs63750066: 6 individuals with 1 copies of risk allele T | *APP* rs63750066 allele T is associated with familial AD [2,3]. |
| *PSEN1* | rs63750306<br>rs63750590<br>rs63750526<br>rs63751235<br>rs661<br>rs63751037<br>rs63749885<br>rs63750231<br>rs63751229<br>rs63751272<br>rs63751223<br>rs63750391<br>rs63751163<br>rs63749891<br>rs281875357<br>rs63751141<br>rs63750082<br>rs121917807<br>rs63750265<br>rs63751144<br>rs63750886<br>rs121917808<br>rs63750599<br>rs63750083<br>rs63749824<br>rs63750577<br>rs267606983<br>rs63750687 | rs63750526<br>rs63749824 | None | rs63749824: 5 individuals with 1 copy of risk allele T<br>rs63750526: 1 individual with 1 copy of risk allele A | *PSEN1* rs63749824 allele T is associated with late-onset AD in a carrier; unaffected mutation carrier has increased CSF Aβ [4].<br>Variant rs63750526 allele A is associated with early-onset AD [5]. |

| | | | | | |
|---|---|---|---|---|---|
| *PSEN2* | rs63750215<br>rs28936379<br>rs63750110<br>rs63750666<br>rs63749851<br>rs63749884<br>rs28936380<br>rs63750197<br>rs63750048 | rs63750110<br>rs63750666<br>rs63750197<br>rs63750048 | None | rs63750048: 1 individual with 1 copy of risk allele T<br>rs63750197: 427 individuals with 1 copy of risk allele T<br>rs63750666: 4 individuals with 1 copy of risk allele T<br>rs63750110: 55 individuals with 1 copy of risk allele C | *PSEN2* rs63750048 allele T is associated with familial AD as well as parkinsonism and Lewy body dementia [6]. Variant rs63750197 was reported in association with AD [7], but ClinVar shows that allele T is benign. Variant rs63750110 allele C is associated with early onset AD in an individual with APOE2/3 genotype [8]. Variant rs63750110 allele C is associated with Alzheimer's disease [8]. |
| *GRN* | rs63750077<br>rs63751006<br>rs63750331<br>rs606231220<br>rs63749801<br>rs606231221<br>rs63751243<br>rs63751294<br>rs63749905<br>rs63751085<br>rs63749877<br>rs5848 | rs63751294<br>rs5848 | rs5848: 4 individuals with 1 copy of risk allele T; 1 individual with 2 copies of risk allele T | rs63751294: 5 individuals with 1 copy of risk allele T<br>rs5848: 2934 individuals with 2 copies of risk allele T | *GRN* rs63751294 allele T is associated with ubiquitin-positive frontotemporal lobar degeneration (FTLD) [9,10]. Variant rs5848 showed significant increase in T/T genotype among FTLD [11]. This variant was only found in the 140k WGS data. |
| *SORT1* | rs12740374 | rs12740374<br>rs17646665 | rs12740374: 1 individual with 1 copy of risk allele T; 2 individuals with 2 copies of risk allele T<br>rs17646665: 1 individual with 1 copy of protective allele G | rs12740374: 13668 individuals with 1 copy of risk allele T; 1927 individuals with 2 copies of risk allele T<br>rs17646665: 2702 individuals with 1 copy of protective allele G; 1926 individuals with 2 copies of protective allele G | *SORT1* rs12740374 allele T creates a transcription factor binding site and results increased SORT1 expression [12], which in turn controls PGRN levels and plays a role in the lysosomal pathway [13]. Variant rs17646665 allele G is significantly associated with reduced risk of AD [14]. |
| *MAPT* | rs63751273<br>rs63750376<br>rs63750424<br>rs63750972<br>rs63750308<br>rs63751011<br>rs1568327531<br>rs63750570 | rs63750424<br>rs63751011<br>rs63750512<br>rs63750129<br>rs63750959<br>rs63750425 | None | rs63751011: 2 individuals with 1 copy of risk allele T<br>rs63750424: 6 individual with 1 copy of risk allele T | *MAPT* rs63751011 allele T is associated with frontotemporal dementia [15-20]. Variant rs63750424 has been associated with frontotemporal dementia [15,21] and has |

| | | | | | |
|---|---|---|---|---|---|
| | rs63750756<br>rs63751165<br>rs63750512<br>rs63751438<br>rs63750912<br>rs63750711<br>rs63750129<br>rs63751264<br>rs63750959<br>rs63750635<br>rs63751394<br>rs63751392<br>rs63750349<br>rs63750425<br>rs63750092<br>rs63751391 | | | | been reported to clinically resembles AD [22]. |
| *APBB2* | rs13133980 | rs13133980 | rs13133980: 2 individuals with 1 copy of risk allele G; 3 individuals with 2 copies of risk allele G | rs13133980: 19,227 individuals with 1 copy of risk allele G; 7,258 individuals with 2 copies of risk allele G | *APBB2* rs13133980 allele G showed stronger association with age of AD onset before 75 years of age [23]. However, another study did not replicate this finding and rs13133980 was not associated with AD regardless of age of onset or APOE status [24]. |

eTable 3. List of self-reported cardiovascular conditions included for analyses

| *Data Field/Table* | *Category* | *Code* | *Description* |
|---|---|---|---|
| *20002* | Self-reported cardiovascular conditions | 1065 | hypertension |
| | | 1066 | heart/cardiac problem |
| | | 1072 | essential hypertension |
| | | 1074 | angina |
| | | 1075 | heart attack/myocardial infarction |
| | | 1076 | heart failure/pulmonary odema |
| | | 1077 | heart arrhythmia |
| | | 1471 | atrial fibrillation |
| | | 1483 | atrial flutter |
| | | 1484 | wolff 10endroflu white / wpw syndrome |
| | | 1485 | irregular heart beat |
| | | 1486 | sick sinus syndrome |
| | | 1487 | svt / supraventricular tachycardia |
| | | 1078 | heart valve problem/heart murmur |
| | | 1584 | mitral valve disease |
| | | 1488 | mitral valve prolapse |
| | | 1489 | mitral stenosis |
| | | 1585 | mitral regurgitation / incompetence |
| | | 1586 | aortic valve disease |
| | | 1490 | aortic stenosis |
| | | 1587 | aortic regurgitation / incompetence |
| | | 1079 | cardiomyopathy |
| | | 1588 | hypertrophic cardiomyopathy (hcm / hocm) |
| | | 1080 | pericardial problem |
| | | 1589 | pericarditis |
| | | 1590 | pericardial effusion |
| | | 1426 | myocarditis |
| | | 1479 | rheumatic fever |
| | | 1081 | stroke |
| | | 1086 | subarachnoid haemorrhage |
| | | 1491 | brain haemorrhage |
| | | 1583 | ischaemic stroke |
| | | 1082 | transient ischaemic attack (tia) |
| | | 1083 | subdural haemorrhage/haematoma |
| | | 1425 | cerebral aneurysm |
| | | 1067 | peripheral vascular disease |
| | | 1087 | leg claudication/ intermittent claudication |

| | | |
|---|---|---|
| | 1088 | arterial embolism |
| | 1492 | aortic aneurysm |
| | 1591 | aortic aneurysm rupture |
| | 1592 | aortic dissection |
| | 1068 | venous thromboembolic disease |
| | 1093 | pulmonary embolism +/- dvt |
| | 1094 | deep venous thrombosis (dvt) |
| | 1473 | high cholesterol |
| | 1493 | other venous/lymphatic disease |
| | 1494 | varicose veins |
| | 1495 | lymphoedema |
| | 1593 | varicose ulcer |

eTable 4. List of self-reported and primary care prescription medications used for analyses

| Data Field/Table | Category | Code | Description |
|---|---|---|---|
| 20003 | Self-reported statins | 1140861958 | simvastatin |
| | | 1140869130 | ecostatin 150mg pessary |
| | | 1140869132 | ecostatin twinpack |
| | | 1140869196 | ecostatin-1 150mg pessary |
| | | 1140870208 | sandostatin 50micrograms/1ml injection |
| | | 1140873350 | imipenem + cilastatin |
| | | 1140873570 | tetracycline+nystatin 250mg/250ku tablet |
| | | 1140874030 | metronidazole+nystatin 400mg/10000units tablet+pessary |
| | | 1140874266 | nystatin dome 100,000units/ml oral suspension |
| | | 1140874360 | nystatin |
| | | 1140878594 | ecostatin cream |
| | | 1140878598 | ecostatin lotion |
| | | 1140880388 | nystatin+tolnaftate 100000units/1%/g cream |
| | | 1140880390 | nystatin+chlorhexidine hydrochloride 100000units/1%/g cream |
| | | 1140882794 | clobetasol propionate+neomycin sulphate+nystatin |
| | | 1140882806 | clobetasone butyrate+oxytetracycline+nystatin |
| | | 1140882844 | hydrocortisone+nystatin |
| | | 1140882938 | terra-cortril nystatin cream |
| | | 1140883060 | triamcinolone+nystatin |
| | | 1140884216 | ecostatin 1% powder |
| | | 1140888594 | fluvastatin |
| | | 1140888648 | pravastatin |
| | | 1140910632 | eptastatin |
| | | 1140910654 | velastatin |
| | | 1141146234 | atorvastatin |
| | | 1141157400 | nystatin product |
| | | 1141192410 | rosuvastatin |
| 20003 | Self-reported non-statin lipid lowering medications | 1140861924 | bezafibrate |
| | | 1140861942 | cholestyramine+aspartame 4g/sachet powder |
| | | 1140861944 | clofibrate |
| | | 1140861954 | fenofibrate |
| | | 1140862026 | ciprofibrate |
| | | 1140865576 | cholestyramine |
| | | 1140888590 | colestipol |
| | | 1140910670 | niacin |
| | | 1141157260 | bezafibrate product |
| | | 1141157262 | gemfibrozil product |

| | | | |
|---|---|---|---|
| | | 1141157416 | cholestyramine product |
| | | 1141192736 | ezetimibe |
| *20003* | Self-reported anti-hypertensive | 1140860192 | nadolol |
| | | 1140860292 | pindolol |
| | | 1140860308 | metoprolol tartrate+chlorthalidone 100mg/12.5mg tablet |
| | | 1140860312 | nadolol+bendrofluazide 40mg/5mg tablet |
| | | 1140860316 | nadolol+bendrofluazide 80mg/5mg tablet |
| | | 1140860322 | pindolol+clopamide 10mg/5mg tablet |
| | | 1140860332 | sotalol hydrochloride+hydrochlorothiazide 80mg/12.5mg tablet |
| | | 1140860336 | timolol maleate+co-amilozide 10mg/2.5mg/25mg tablet |
| | | 1140860340 | timolol maleate+bendrofluazide 10mg/2.5mg tablet |
| | | 1140860342 | timolol maleate+bendrofluazide 20mg/5mg tablet |
| | | 1140860404 | metoprolol tartrate+hydrochlorothiazide 100mg/12.5mg tablet |
| | | 1140860418 | propranolol hydrochloride+bendrofluazide 80mg/2.5mg capsule |
| | | 1140860422 | acebutolol+hydrochlorothiazide 200mg/12.5mg tablet |
| | | 1140860426 | atenolol+nifedipine 50mg/20mg m/r capsule |
| | | 1140860470 | methyldopa |
| | | 1140860532 | minoxidil |
| | | 1140860562 | methyldopa+hydrochlorothiazide 250mg/15mg tablet |
| | | 1140860696 | lisinopril |
| | | 1140860728 | quinapril |
| | | 1140860738 | quinalapril+hydrochlorothiazide 10mg/12.5mg tablet |
| | | 1140860750 | captopril |
| | | 1140860764 | captopril+hydrochlorothiazide 25mg/12.5mg tablet |
| | | 1140860790 | enalapril maleate+hydrochlorothiazide 20mg/12.5mg tablet |
| | | 1140860806 | ramipril |
| | | 1140860904 | trandolapril |
| | | 1140861088 | nifedipine |
| | | 1140861190 | isradipine |
| | | 1140864202 | chlorthalidone tablet+potassium m/r tablet 25mg/6.7mmol pack |
| | | 1140864950 | bisoprolol fumarate+hydrochlorothiazide 10mg/6.25mg tablet |
| | | 1140864952 | lisinopril+hydrochlorothiazide 10mg/12.5mg tablet |
| | | 1140866078 | indapamide |
| | | 1140866092 | metolazone |
| | | 1140866138 | chlorothiazide |
| | | 1140866144 | chlorthalidone |
| | | 1140866162 | hydrochlorothiazide |
| | | 1140866236 | spironolactone |

| | |
|---|---|
| 1140866280 | bumetanide |
| 1140866324 | triamterene+benzthiazide 50mg/25mg capsule |
| 1140866330 | triamterene+chlorthalidone 50mg/50mg tablet |
| 1140866332 | triamterene+frusemide 50mg/40mg tablet |
| 1140866388 | triamterene |
| 1140866422 | amiloride hcl+cyclopenthiazide 2.5mg/250micrograms tablet |
| 1140866426 | amiloride hydrochloride+bumetanide 5mg/1mg tablet |
| 1140866448 | bumetanide+potassium 500micrograms/7.7mmol m/r tablet |
| 1140866724 | acebutolol |
| 1140866738 | atenolol |
| 1140871986 | clonidine hydrochloride 25micrograms tablet |
| 1140875840 | timolol 0.25% eye drops |
| 1140875934 | apraclonidine |
| 1140879758 | betaxolol |
| 1140879760 | bisoprolol |
| 1140879778 | doxazosin |
| 1140879794 | prazosin |
| 1140879798 | terazosin |
| 1140879802 | amlodipine |
| 1140879806 | diltiazem |
| 1140879810 | nicardipine |
| 1140879824 | labetalol |
| 1140879842 | propranolol |
| 1140879866 | timolol |
| 1140883468 | clonidine |
| 1140888510 | verapamil |
| 1140888512 | amiloride |
| 1140888552 | enalapril |
| 1140888556 | fosinopril |
| 1140888560 | perindopril |
| 1140888646 | felodipine |
| 1140888686 | hydralazine |
| 1140909368 | carvedilol |
| 1140909708 | furosemide |
| 1140910606 | alpha methyldopa |
| 1140916356 | losartan |
| 1140923712 | moexipril |
| 1140926778 | diltiazem hcl+hydrochlorothiazide 150mg/12.5mg m/r capsule |
| 1140928226 | nisoldipine |
| 1141145660 | valsartan |

| | | | |
|---|---|---|---|
| | | 1141146124 | atenolol+chlorthalidone |
| | | 1141146126 | atenolol+bendrofluazide |
| | | 1141146128 | atenolol+co-amilozide |
| | | 1141151016 | losartan potassium+hydrochlorothiazide 50mg/12.5mg tablet |
| | | 1141152998 | irbesartan |
| | | 1141153328 | trandolapril+verapamil hydrochloride |
| | | 1141156836 | candesartan cilexetil |
| | | 1141165470 | felodipine+ramipril |
| | | 1141166006 | telmisartan |
| | | 1141169516 | dorzolamide+timolol |
| | | 1141171336 | eprosartan |
| | | 1141172682 | irbesartan+hydrochlorothiazide 150mg/12.5mg tablet |
| | | 1141180592 | perindopril+indapamide |
| | | 1141180772 | triamterene+chlortalidone 50mg/50mg tablet |
| | | 1141180778 | atenolol+chlortalidone |
| | | 1141184722 | latanoprost+timolol |
| | | 1141187788 | telmisartan+hydrochlorothiazide 40mg/12.5mg tablet |
| | | 1141194804 | nadolol+15endroflumethiazide 40mg/5mg tablet |
| | | 1141194808 | timolol maleate+15endroflumethiazide 10mg/2.5mg tablet |
| | | 1141194810 | atenolol+bendroflumethiazide |
| | | 1141195254 | triamterene+furosemide 50mg/40mg tablet |
| | | 1141195258 | furosemide+potassium 20mg/10mmol m/r tablet |
| | | 1141201038 | valsartan+hydrochlorothiazide 80mg/12.5mg tablet |
| | | 1141201244 | Eplerenone |
| *gp_scripts* | Primary care prescription for lipid modifying agents | C10AA | HMG CoA reductase inhibitors (statins) |
| | | C10AB | Fibrates |
| | | C10AC | Bile acid sequestrants |
| | | C10AD | Nicotinic acid and derivatives |
| | | C10AX | Other lipid modifying agents |
| | | C10BA | Combinations of various lipid modifying agents |
| | | C10BX | Lipid modifying agents in combination with other drugs |
| *gp_scripts* | Primary care prescription for antihypertensives | C02A | Antiadrenergic agents, centrally acting |
| | | C02B | Antiadrenergic agents, ganglion-blocking |
| | | C02C | Antiadrenergic agents, peripherally acting |
| | | C02D | Arteriolar smooth muscle, agents acting on |
| | | C02K | Other antihypertensives |
| | | C02L | Antihypertensives and diuretics in combination |
| | | C02N | Combinations of antihypertensives in ATC-GR. C02 |
| | | C03A | Low-ceiling diuretics, thiazides |
| *gp_scripts* | | C03B | Low-ceiling diuretics, excl. thiazides |

| | | | |
|---|---|---|---|
| | Primary care prescription for diuretics | C03C | High-ceiling diuretics |
| | | C03D | Aldosterone antagonists and other potassium-sparing agents |
| | | C03E | Diuretics and potassium-sparing agents in combination |
| | | C03X | Other diuretics |
| *gp_scripts* | Primary care prescription for beta blocking agents | C07A | Beta blocking agents |
| | | C07B | Beta blocking agents and thiazides |
| | | C07C | Beta blocking agents and other diuretics |
| | | C07D | Beta blocking agents, thiazides and other diuretics |
| | | C07E | Beta blocking agents and vasodilators |
| | | C07F | Beta blocking agents, other combinations |
| *gp_scripts* | Primary care prescription for calcium channel blockers | C08C | Selective calcium channel blockers with mainly vascular effects |
| | | C08D | Selective calcium channel blockers with direct cardiac effects |
| | | C08E | Non-selective calcium channel blockers |
| | | C08G | Calcium channel blockers and diuretics |
| *gp_scripts* | Primary care prescription for agents acting on the renin-angiotensin system | C09A | ACE inhibitor, plain |
| | | C09B | ACE inhibitor, combinations |
| | | C09C | Angiotensin II receptor blockers (ARBs), plain |
| | | C09D | Angiotensin II receptor blockers (ARBs), combinations |
| | | C09X | Other agents acting on the renin-angiotensin system |

eTable 5. Availability and overlap of APOECh carriers with brain imaging, metabolomics, and proteomics data

| | *Brain MRI completed* | *Metabolomics* | *Proteomics* |
|---|---|---|---|
| *Brain MRI completed* | 3 | 1 | 1 |
| *Metabolomics* | | 9 | 2 |
| *Proteomics* | | | 7 |

eTable 6. List of quantitative traits assessed for carriers and noncarriers

| *Category* | *UK Biobank Data Field* | *Description* |
|---|---|---|
| *Physical measures* | 48 | Waist circumference |
| | 49 | Hip circumference |
| | 4079 | Diastolic Blood Pressure, automated reading |
| | 4080 | Systolic Blood Pressure, automated reading |
| | 21001 | Body mass index (BMI) |
| | 21002 | Weight |
| | 21021 | Pulse wave Arterial Stiffness index |
| *Blood biomarkers* | 30600 | Albumin |
| | 30610 | Alkaline phosphatase |
| | 30620 | Alanine aminotransferase |
| | 30630 | Apolipoprotein A |
| | 30640 | Apolipoprotein B |
| | 30650 | Aspartate aminotransferase |
| | 30660 | Direct bilirubin |
| | 30670 | Urea |
| | 30680 | Calcium |
| | 30690 | Cholesterol |
| | 30700 | Creatinine |
| | 30710 | C-reactive protein |
| | 30720 | Cystatin C |
| | 30730 | Gamma glutamyltransferase |
| | 30740 | Glucose |
| | 30750 | Glycated haemoglobin (HbA1c) |
| | 30760 | HDL cholesterol |
| | 30770 | IGF-1 |
| | 30780 | LDL direct |
| | 30790 | Lipoprotein A |
| | 30800 | Oestradiol |
| | 30810 | Phosphate |
| | 30820 | Rheumatoid factor |
| | 30830 | SHBG |
| | 30840 | Total bilirubin |
| | 30850 | Testosterone |
| | 30860 | Total protein |
| | 30870 | Triglycerides |
| | 30880 | Urate |
| | 30890 | Vitamin D |
| *Hematological traits* | 30000 | White blood cell (leukocyte) count |
| | 30010 | Red blood cell (erythrocyte) count |
| | 30020 | Haemoglobin concentration |
| | 30030 | Haematocrit percentage |
| | 30040 | Mean corpuscular volume |
| | 30050 | Mean corpuscular haemoglobin |
| | 30060 | Mean corpuscular haemoglobin concentration |
| | 30070 | Red blood cell (erythrocyte) distribution width |
| | 30080 | Platelet count |
| | 30090 | Platelet crit |
| | 30100 | Mean platelet (thrombocyte) volume |
| | 30110 | Platelet distribution width |

| | | |
|---|---|---|
| | 30120 | Lymphocyte count |
| | 30130 | Monocyte count |
| | 30140 | Neutrophill count |
| | 30150 | Eosinophill count |
| | 30160 | Basophill count |
| | 30170 | Nucleated red blood cell count |
| | 30180 | Lymphocyte percentage |
| | 30190 | Monocyte percentage |
| | 30200 | Neutrophill percentage |
| | 30210 | Eosinophill percentage |
| | 30220 | Basophill percentage |
| | 30230 | Nucleated red blood cell percentage |
| | 30240 | Reticulocyte percentage |
| | 30250 | Reticulocyte count |
| | 30260 | Mean reticulocyte volume |
| | 30270 | Mean sphered cell volume |
| | 30280 | Immature reticulocyte fraction |
| | 30290 | High light scatter reticulocyte percentage |
| | 30300 | High light scatter reticulocyte count |
| *Urine biomarkers* | 30500 | Microalbumin in urine |
| | 30510 | Creatinine (enzymatic) in urine |
| | 30520 | Potassium in urine |
| | 30530 | Sodium in urine |
| *Cognition* | 20016 | Fluid intelligence score (assessment center touchscreen) |
| | 20023 | Reaction time: Mean time to correctly identify matches |
| | 6138, 10722 | Educational qualifications, education qualifications pilot |

eTable 7. Empirical p-values of binary traits

| *Trait* | *APOECh carriers with event (N=36)* | *Noncarriers with event (N=129,240)* | *P-value* |
|---|---|---|---|
| *AD* | 0 | 660 (0.51%) | 0.83 |
| *Parental history of AD* | 4 (11.1%) | 17428 (13.5%) | 0.74 |
| *MCI or other cognitive function symptoms* | 0 | 755 (0.58%) | 0.81 |
| *CVD (ICD + self-report)* | 22 (61.1%) | 72652 (56.2%) | 0.34 |
| *Dyslipidemia (ICD + self-report)* | 10 (27.8%) | 24643 (19.1%) | 0.13 |
| *Primary hypertension (ICD + self-report)* | 15 (41.7%) | 44869 (34.7%) | 0.24 |
| *Self-reported statin at baseline* | 5 (13.9%) | 15970 (12.3%) | 0.46 |
| *Prescription for lipid-lowering drugs* | 6 (35.3%) | 16436 (12.7%) | 0.30 |
| *Use of lipid-lowering drugs (prescription + self-report)* | 9 (25%) | 27281 (21.1%) | 0.34 |
| *Prescription for antihypertensive drugs* | 9 (52.9%) | 22011 (17.0%) | 0.15 |
| *Use of antihypertensive (prescription + self-report)* | 12 (33.3%) | 36029 (27.9%) | 0.29 |

eTable 8. Linear and logistic regression of HDL, LDL, CVD, and AD with PRS as a covariate between APOECh carriers and noncarriers

| *Trait* | *PRS UKB Data Field* | *Cases in carriers (N=36)* | *Cases in noncarriers (N=129,240)* | *Slope of carrier status* | *P-value of carrier status* |
|---|---|---|---|---|---|
| *HDL* | 26242 | NA | NA | -0.03 | 0.64 |
| *LDL* | 26250 | NA | NA | -0.21 | 0.16 |
| *CVD* | 26223 | 22 | 72,651 | 0.22 | 0.52 |
| *AD* | 26206 | 0 | 660 | 0.69 | 0.95 |

eTable 9. Kolmogorov-Smirnov test of quantitative traits and PRS among all 36 carriers and matching controls

| *Category* | *UKB Data Field* | *Median of carriers N=36* | *Median of noncarriers N=129,240* | *KS test* |
|---|---|---|---|---|
| *Lipid biomarkers* | Apolipoprotein_A_30630 | 1.64 | 1.55 | D=0.19 P-value=0.239 |
| | Apolipoprotein_B_30640 | 0.88 | 1.02 | D=0.31 P-value=0.004 |
| | statin_adj_Apolipoprotein_B_30640 | 0.94 | 1.06 | D=0.25 P-value=0.036 |
| | C-Reactive_protein_30710 | 1.12 | 1.32 | D=0.12 P-value=0.662 |
| | Cholesterol_30690 | 5.32 | 5.73 | D=0.18 P-value=0.263 |
| | statin_adj_Cholesterol_30690 | 5.80 | 5.91 | D=0.15 P-value=0.427 |
| | HDL_30760 | 1.56 | 1.47 | D=0.2 P-value=0.225 |
| | LDL_30780 | 3.25 | 3.55 | D=0.23 P-value=0.072 |
| | statin_adj_LDL_30780 | 3.36 | 3.70 | D=0.21 P-value=0.119 |
| | Lipoprotein_A_30790 | 22.00 | 20.50 | D=0.11 P-value=0.838 |
| | Triglyceride_30870 | 1.71 | 1.42 | D=0.16 P-value=0.329 |
| *Physical measures* | DBP_4079 | 83.00 | 81.00 | D=0.1 P-value=0.726 |
| | adjusted_DBP_4079 | 83.25 | 83.00 | D=0.1 P-value=0.806 |
| | SBP_4080 | 129.50 | 136.00 | D=0.15 P-value=0.332 |
| | adjusted_SBP_4080 | 136.25 | 137.00 | D=0.11 P-value=0.755 |
| | Pulse_wave_Arterial_Stiffness_index_21021 | 10.19 | 8.78 | D=0.38 P-value=0.149 |
| | Waist_circumference_48 | 86.00 | 87.00 | D=0.1 P-value=0.694 |
| | Hip_circumference_49 | 102.00 | 102.00 | D=0.13 P-value=0.411 |
| | Body_mass_index_21001 | 25.65 | 26.47 | D=0.16 P-value=0.3 |
| | Weight_21002 | 71.70 | 73.50 | D=0.15 P-value=0.375 |
| *Urine biomarkers* | Microalbumin_30500 | 7.80 | 11.20 | D=0.54 P-value=0.02 |
| | Creatinine_30510 | 8737.00 | 6792.00 | D=0.14 P-value=0.48 |
| | Potassium_30520 | 58.60 | 55.00 | D=0.08 P-value=0.957 |
| | Sodium_30530 | 60.20 | 63.70 | D=0.08 P-value=0.972 |
| *Blood biomarkers* | Albumin_30600 | 44.86 | 45.14 | D=0.16 P-value=0.444 |
| | Alkaline_phosphatase_30610 | 84.10 | 80.30 | D=0.16 P-value=0.327 |
| | Alanine_aminotransferase_30620 | 20.52 | 18.94 | D=0.14 P-value=0.516 |
| | Aspartate_aminotransferase_30650 | 26.70 | 23.70 | D=0.25 P-value=0.029 |
| | Direct_bilirubin_30660 | 1.42 | 1.55 | D=0.16 P-value=0.411 |
| | Urea_30670 | 4.90 | 5.17 | D=0.12 P-value=0.702 |
| | Calcium_30680 | 2.39 | 2.38 | D=0.16 P-value=0.494 |
| | Creatinine_30700 | 69.90 | 66.90 | D=0.17 P-value=0.288 |
| | Cystatin_C_30720 | 0.86 | 0.87 | D=0.1 P-value=0.887 |
| | Gamma_glutamyltransferase_30730 | 25.60 | 23.90 | D=0.13 P-value=0.624 |
| | Glucose_30740 | 4.71 | 4.91 | D=0.25 P-value=0.067 |
| | HbA1c_30750 | 35.00 | 35.00 | D=0.12 P-value=0.609 |
| | IGF-1_30770 | 20.47 | 21.17 | D=0.17 P-value=0.29 |
| | Oestradiol_30800 | 396.40 | 366.20 | D=0.26 P-value=0.585 |
| | Phosphate_30810 | 1.22 | 1.17 | D=0.13 P-value=0.685 |
| | Rheumatoid_factor_30820 | 12.65 | 16.50 | D=0.65 P-value=0.239 |
| | SHBG_30830 | 49.88 | 49.66 | D=0.12 P-value=0.821 |
| | Total_bilirubin_30840 | 7.60 | 7.70 | D=0.07 P-value=0.996 |
| | Testosterone_30850 | 1.68 | 1.34 | D=0.13 P-value=0.72 |
| | Total_protein_30860 | 71.92 | 72.15 | D=0.12 P-value=0.777 |
| | Urate_30880 | 303.40 | 284.20 | D=0.16 P-value=0.338 |

| | | | | |
|---|---|---|---|---|
| | Vitamin D_30890 | 56.80 | 47.30 | D=0.21 P-value=0.115 |
| *Hematological traits* | Leukocyte_count_30000 | 6.36 | 6.65 | D=0.12 P-value=0.691 |
| | Erythrocyte_count_30010 | 4.50 | 4.41 | D=0.13 P-value=0.543 |
| | Haemoglobin_concentration_30020 | 13.70 | 13.86 | D=0.12 P-value=0.645 |
| | Haematrocrit_percentage_30030 | 40.51 | 40.20 | D=0.11 P-value=0.807 |
| | Mean_corpuscular_volume_30040 | 90.85 | 91.29 | D=0.07 P-value=0.995 |
| | Mean_corpuscular_haemoglobin_30050 | 31.47 | 31.50 | D=0.12 P-value=0.675 |
| | Mean_corpuscular_haemoglobin_concentration_30060 | 34.19 | 34.45 | D=0.15 P-value=0.404 |
| | Erythrocyte_distribution width_30070 | 13.50 | 13.31 | D=0.23 P-value=0.038 |
| | Platelet_count_30080 | 255.00 | 254.00 | D=0.11 P-value=0.78 |
| | Platelet crit_30090 | 0.25 | 0.24 | D=0.18 P-value=0.195 |
| | Thrombocyte_volume_30100 | 9.60 | 9.20 | D=0.26 P-value=0.017 |
| | Platelet_distribution_width_30110 | 16.48 | 16.40 | D=0.13 P-value=0.537 |
| | Lymphocyte_count_30120 | 1.80 | 1.89 | D=0.22 P-value=0.067 |
| | Monocyte_count_30130 | 0.42 | 0.43 | D=0.08 P-value=0.924 |
| | Neutrophill_count_30140 | 4.09 | 4.03 | D=0.09 P-value=0.891 |
| | Eosinophill_count_30150 | 0.13 | 0.13 | D=0.09 P-value=0.814 |
| | Basophill_count_30160 | 0.02 | 0.02 | D=0.1 P-value=0.599 |
| | Nucleated_red_blood_cell_count_30170 | 0.00 | 0.00 | D=0.01 P-value=1 |
| | Lymphocyte_percentage_30180 | 26.20 | 28.68 | D=0.19 P-value=0.142 |
| | Monocyte_percentage_30190 | 6.48 | 6.68 | D=0.13 P-value=0.615 |
| | Neutrophill_percentage_30200 | 61.70 | 61.30 | D=0.18 P-value=0.18 |
| | Eosinophill_percentage_30210 | 1.80 | 2.08 | D=0.1 P-value=0.841 |
| | Basophill_percentage_30220 | 0.40 | 0.43 | D=0.14 P-value=0.466 |
| | Nucleated_red_blood_cell_percentage_30230 | 0.00 | 0.00 | D=0.01 P-value=1 |
| | Reticulocyte_percentage_30240 | 1.09 | 1.25 | D=0.2 P-value=0.126 |
| | Reticulocyte_count_30250 | 0.05 | 0.06 | D=0.18 P-value=0.183 |
| | Mean_reticulocyte_volume_30260 | 107.42 | 105.61 | D=0.14 P-value=0.451 |
| | Mean_sphered_cell_volume_30270 | 84.30 | 82.70 | D=0.16 P-value=0.333 |
| | Immature_reticulocyte_fraction_30280 | 0.27 | 0.29 | D=0.22 P-value=0.053 |
| | High_light_scatter_reticulocyte_percentage_30290 | 0.27 | 0.36 | D=0.22 P-value=0.073 |
| | High_light_scatter_reticulocyte_count_30300 | 0.01 | 0.02 | D=0.23 P-value=0.03 |
| *Cognition* | education_years | 6 | 6 | D=0.24 P-value=0.316 |
| | fluid_intelligence_score_center_20016 | 522 | 535 | D=0.07 P-value=0.98 |
| | reaction_time_20023 | 14 | 15 | D=0.11 P-value=0.436 |
| *Dyslipidemia PRS* | HDL_26242 | -0.04 | 0.01 | D=0.13 P-value=0.53 |
| | LDL_26250 | -0.11 | -0.06 | D=0.1 P-value=0.79 |
| *Cardiovascular PRS* | Atrial_fibrillation_26212 | 0.33 | 0.10 | D=0.12 P-value=0.58 |
| | Cardiovascular_disease_26223 | -0.35 | -0.10 | D=0.16 P-value=0.29 |
| | Coronary_artery_disease_26227 | -0.31 | -0.16 | D=0.14 P-value=0.48 |
| | Hypertension_26244 | 0.09 | -0.04 | D=0.15 P-value=0.34 |
| | Stroke_26248 | -0.04 | -0.02 | D=0.08 P-value=0.96 |
| *Neurological disease PRS* | Alzheimer's_disease_26206 | -0.31 | -0.08 | D=0.25 P-value=0.02 |
| | Multiple_sclerosis_26254 | 0.06 | -0.15 | D=0.15 P-value=0.33 |
| | Parkinson's_disease_26260 | -0.54 | -0.17 | D=0.21 P-value=0.06 |

eTable 10. Summary of phenotypic data availability for carriers and noncarriers

| | *All APOECh carriers (N=37)* | *APOECh EUR carriers (N=36)* | *Matched EUR noncarriers (N=129,240)* | *P-value for differences in EUR data availability* |
|---|---|---|---|---|
| *≥ 1 ICD-9 or ICD-10 code* | 30 (81.1%) | 30 (83.3%) | 113,386 (87.7%) | 0.85 |
| *≥ 1 Self-reported non-cancer illness(es)* | 24 (64.7%) | 23 (63.9%) | 98,938 (76.6%) | 0.97 |
| *≥ 1 Self-reported medication(s)* | 30 (81.1%) | 29 (80.6%) | 97,146 (75.2%) | 0.30 |
| *Primary care data available* | 18 (48.6%) | 17 (47.2%) | 60,680 (46.9%) | 0.55 |
| *Primary care prescription data available* | 18 (48.6%) | 17 (47.2%) | 58,575 (45.3%) | 0.47 |
| *≥ 1 lipid biomarker measurement* | 32 (86.5%) | 31 (86.1%) | 123,587 (95.6%) | 0.995 |
